# Clinical Impact of *IDH1* Mutations and *MGMT* Methylation in Adult Glioblastoma Multiforme

**DOI:** 10.1101/2022.03.30.22273163

**Authors:** Magda Sayed Mahmoud, Mohamed K. Khalifa, Amira M Nageeb, Lobna Ezz El-Arab, Manal El-Mahdy, Amal Ramadan, Maha Hashim, Noha Bakr, Menha Swellam

## Abstract

**Background:** Genetic aberrations and epigenetic alterations have been reported in different types of cancer. Impact of Isocitrate dehydrogenase1 (*IDH1*) and O6-methylguanine-DNAmethyltransferase (*MGMT*) in glioblastoma multiforme (GBM) have been of great interest due to their implications in prediction of prognosis of several types of cancer. Authors aimed to investigate the clinical role of *IDH1* mutation and *MGMT* methylation pattern among GBM patients versus non-neurooncological diseases (NND) patients and their impact on survival criteria.

**Methods:** Formalin-Fixed Paraffin-Embedded (FFPE) tissue sections of 58 GBM and 20 non-onconeurological diseases patients were recruited and *IDH1* mutation were detected using Cast-PCR technology and *MGMT* methylation was detected using Methyl II quantitative PCR approach. Their results were assessed with other clinicopathological criteria and assess its correlation with survival patterns.

**Results:** *IDH1* mutation was detected among 15 GBM cases (15/58) and it was not reported among NND (*P*=0.011). Receiver operating characteristic (ROC) curve were plotted to discriminate between *MGMT* methylation among studied groups. Patients with *MGMT* methylation ≥ 66% was reported as high methylation, which was recorded significantly in 51.7% and 100% of GBM cases and NND, respectively. Both showed significant difference with performance status, while *MGMT* methylation was significantly related with tumor size and tumor location. *IDH1* mutation and *MGMT* methylation reported significant increase with GBM patients revealed complete response to treatment. Survival pattern was better for *IDH1* mutation and *MGMT* high methylation as compared to *IDH1* wild type or *MGMT* low-moderate methylation, respectively and favorable survival was detected when both were combined than using either of them alone.

**Conclusion:** Detection of *IDH1* mutation and *MGMT* methylation among GBM patients could aid in prediction of their response to treatment and their survival patterns, and their combination is better than using any of them alone.

## Background

Glioblastoma Multiforme (GBM) is the prevalent malignant tumor of the brain with poor prognosis and aggressive development without identifiable precursor lesions [1]. Advances have been made in therapeutic strategies after addition of slandered of care chemotherapy treatment i.e. temozolomide (TMZ) to maximal safe tumor resection and radiotherapy. However, median survival is still limited to 15 months [2,3].Thus, there is a great need to unravel oncogenic mechanisms of GBM since there are two types of this kind of malignancy either displayed rapidly de novo with unknown precursor lesion or from low-grade tumor [4] although they cannot be histopathologically distinguished but both with different molecular alterations due to different genes have been reported to be involved in the process of GBM pathogenicity [5].

On the genetic level several alterations have been reported in glioblastoma as alteration in TP53 [6], truncated of activated form of EGFR (i.e. EGFRvIII) [7], deletions in *PTEN* [8] and mutation in *BRCA* genes [9]. Also, by using genome - wide sequencing it has been reported that isocitrate dehydrogenases genes (*IDH1* and *IDH2*) are mutated among GBM patients especially in younger patients and among secondary GBM patients [10] and were independent prognostic marker.

Moreover, GBM presents with a range of epigenetic changes which are mitotically heritable alterations in gene expression that are not related to alterations in DNA sequence [11]. Among the studied epigenetic variations is CpG island hypermethylation which has been reported in several types of cancer [11-13]. O-6-methylguanine-DNA methyltransferase (*MGMT*), one of the DNA repair enzymes which participate in GBM resistance to TMZ by *MGMT* methylation in the promoter region [2]. Silencing of MGMT has been correlated with increased survival independently of treatment strategy [14].

Although both genetic and epigenetic variations in GBM have been studied independently, there is an indication that these mechanisms interact on signaling pathways, chromosomal domains, and certain genes [4].

In the current study, authors aimed to investigate both *IDH1* genetic mutation and *MGMT* epigenetic methylation of *MGMT* among GBM patients and investigate their correlation with each other as well as their relation as predictive prognostic markers among Egyptian patients when tested alone or in combination.

## Subjects and methods

### Patient selection

After getting ethical approval from Medical Ethical Committee of National Research Centre (ID#20110), to recruit patients who fulfilled the inclusion criteria as adult persons (age > 18 years) newly diagnosed GBM with performance < or = 2 according to the Ester Clinical Oncology Group; which assesses disease progression affecting on patient’s daily living abilities and patients with non-neurological diseases and have no other malignancies, while GBM patients who have not fulfilled these criteria were not recruited. Accordingly, GBM patients (n=58) patients were recruited after signing their informed consent. Also a group of non-neurooncological diseases (NND) were recruited (n=20). Tumor tissue samples were surgically resected by stereotactic*/*open biopsy of enrolled brain tumors then fixed in formalin buffer and fixed in paraffin stained with hematoxylin-eosin (HE) reviewed by neuropathologists (MM) to confirm diagnosis according to WHO classification [15]. Then 5-10 sections from FFPE were transferred to Eppendorf tubes for further processing of DNA extraction.

### Extraction DNA

Extraction of DNA from FFPE samples was done using QIAamp FFPE kit (Cat no. 56404) according to manufacturer instructions and both concentration and purity were detected using nono-drop spectrophotometer (Quawell, Q-500, Scribner, USA) at absorbance (260 and 280 nm) and tested on 1% agarose gel, the extracted DNA samples were stored in -20 for further processing to detect *MGMT* and *IDH1* mutation.

### Detection of MGMT methylation pattern using Methyl II quantitative PCR system

MGMT methylation pattern was detected in DNA extracted samples using EpiTect Methyl II quantitative polymerase chain reaction (qPCR) System (Qiagen, Germany) which based on assessment of residual extracted DNA after cleavage with restriction enzyme then the remaining DNA will be quantified by real-time PCR using specific primers for the desired gene that flanks a promoter region of interest. Thus reaction was performed in two phases with some modifications in our lab: phase I: carried out using EpiTect Methyl II DNA Restriction Kit (cat. no. 335452), briefly extracted genomic DNA was aliquoted into two equivalent portions into 2 PCR reaction tubes and they were designated as follows: no-enzyme (UD i.e. no restriction enzyme was added), methylation-sensitive restriction enzyme (D i.e. restriction enzyme sensitive to methylation hence digest unmethylated DNA) then using thermal cycler (SureCycler 8800, Agilent, Santa Clara, CA, USA) they were incubated at 37 °C for 6 h then at 65 °C for 20 min. The remaining genomic DNA sample in each tube (UD and D) is quantified through phase II which was carried out using QPCR (Max3005P; Stratagene, AgilentTechnologies, CA, USA). Briefly; 5ul from the remaining DNA was mixed with qPCR master mix (RT2 qPCRSYBR Green/ROX Master Mix, Cat number 330520) and were distributed into a PCR plate with pre-aliquoted *MGMT* primer as follows Left primer ATTTTTGTGATAGGAAAAGGTATGG Right primer CTAAAACAATCTACACATCCTCACT. QPCR is done using cycling conditions as follows: 95 °C for 10 min (1 cycle), then 99 °C for 30 and 72 °C for 1 min (3 cycles), and finally 97 °C for 15 sec and 72 °C for 1 min (40 cycles). At the end, the raw ΔCT values were collected for each PCR reaction tube (UD and D) for each sample as shown in Figure (1A-B). Because in the qPCR reaction the UD was used hence the DNA in which all CpG sites are methylated will be detected by real-time PCR [16] through following equations:

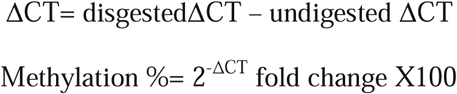

**Figure 1:**
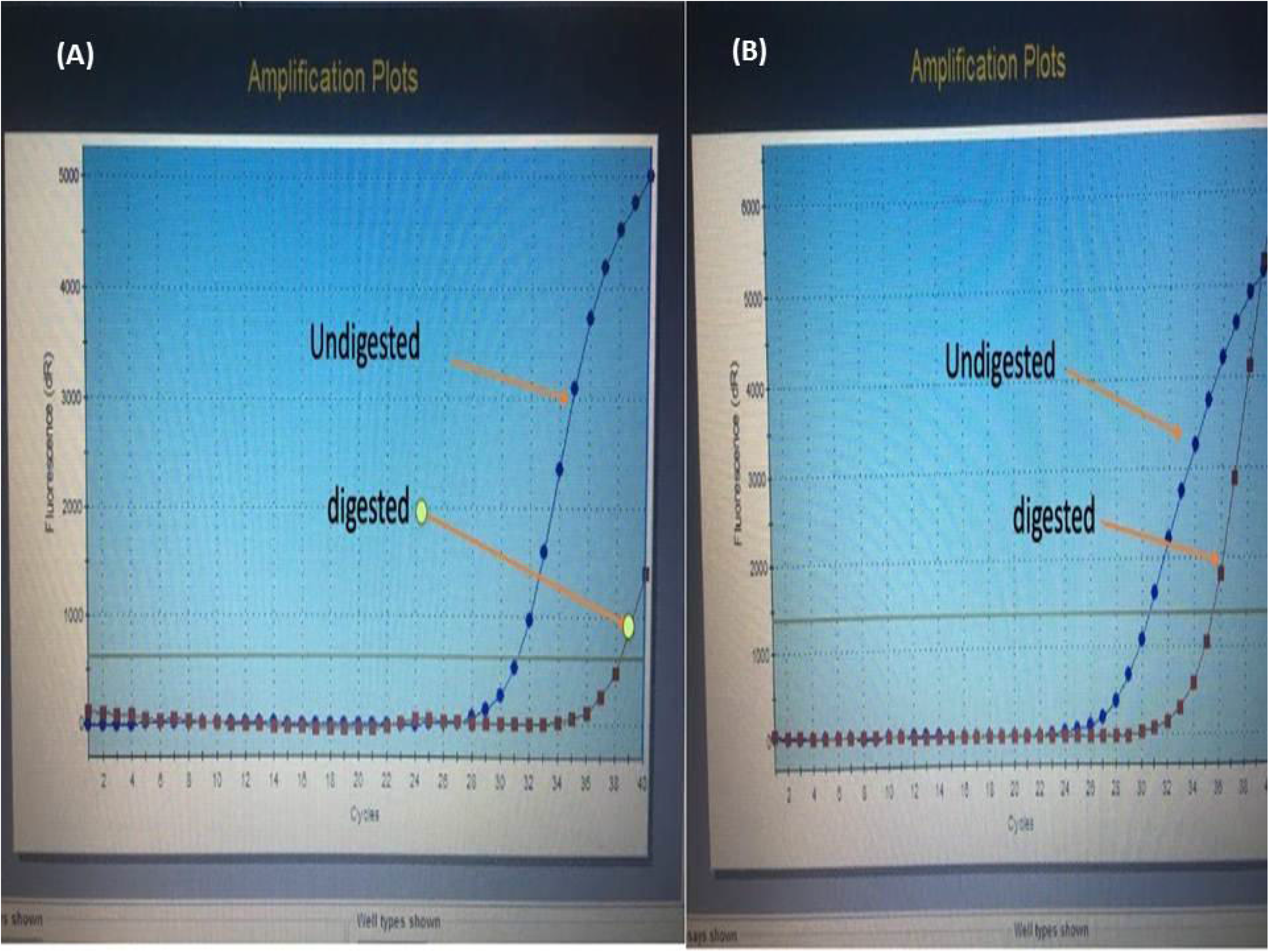
Amplification plots of the *MGMT* qPCR reaction, A: showing plots of 100 % unmethylated sample, and B showing 12.5 % methylated sample.

### Determination of IDH1 mutation using Cast-PCR technology

Competitive allele specific TaqMan PCR (Cast-PCR) technology was used to detect *IDH1* mutation as it is sensitive, specific and fast method for detection of mutant allele since it permits the discriminating amplification of minor alleles and blocks the amplification of non-mutant allele [17,18].Qualitative assessment of 6 mutations within *IDH1* mutation codon 132 (the 2 major R132H and R132C mutations, and 4 “*IDH1*-other”: R132G, R132S, R132L, R132V), one within *IDH1* codon 100 (R100Q), Amplification Refractory Mutation System (ARMS) PCR was combined to identify the most common *IDH1* R132H/R132C. Preparation of master mix was recommended by the supplier. Fifty ng of gDNA per reaction and the described probes above were used. Then PCR were performed as follows: pre-PCR read 60°C for 30 s; holding stage 50°C for 2 min, 95°C for 10 min; cyclingstage 95°C for 15 s, 60°C for 1 min for 40 cycles; and post-PCR 60°C for 30 s. The limit of detection (LOD) of castPCR TM was assessed by constructing dilution curves of samples from patients with and without *IDH1/2* gene mutations. Each point was determined using different dilutions (1:1 to 1:50) of the mutated sample and a non-mutated sample (Figure 2A-B). Sensitivity and specificity of the Cast-PCR for *IDH1* R132H (SNVs) allowing over 99% confidence of detecting down to 5% mutant DNA in a wild-type background.

**Figure 2:**
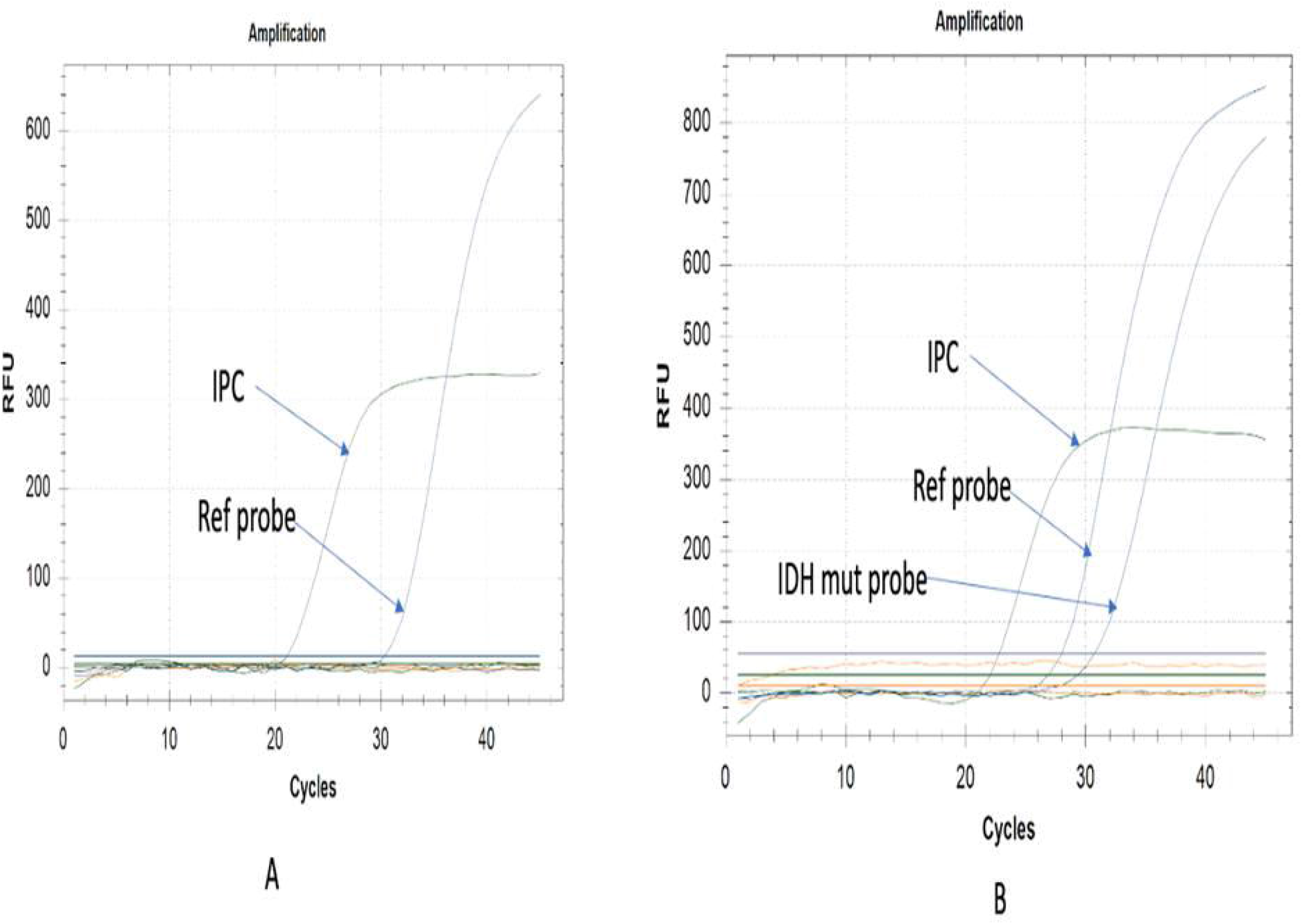
Amplification plots of the *IDH1* mutation by Cast PCR reaction, A: showing plots of wild type sample, and B showing positive mutation sample.

### Pathology Preparation

This study included formalin-fixed paraffin-embedded tissue blocks from patients with glioblastoma. Inclusion criteria for tumor tissue blocks are as follows: (1) histopathological diagnosis of glioblastoma with more than 80% viable tumor tissue, (2) available archival paraffin-embedded tissue blocks, and (3) available clinical follow-up data. All studied glioblastoma cases and non-neoplastic control cases were subjected to the following: I) The paraffin-embedded tissue blocks of the studied glioblastoma cases were cut at full sections with a thickness of 4 microns and stained for routine H&E stain. The stained slides of the tissue specimens have been prepared to confirm the diagnosis based on the 2016 CNS WHO classification and to assess viability of the submitted tumor tissue. II) For preparation of PCR testing, freshly cut sections of paraffin-embedded tissue, each with a thickness of up to 10 um. Up to 8 sections, each with a thickness of up to 10 um and a surface area of up to 250 mm2, can be combined in one preparation.

### Treatment strategies

The Clinical Target Volume (CTV) was contoured on computerized tomography (CT) and postoperative magnetic resonance imaging (MRI) image fusion with integrated residual tumor mass (T1 gadolinium-enhanced lesion) and/or postoperative cavity (i.e., GTV) plus a 15–20mm margin without reflection for peri-tumoral edema. Volume contouring took into account anatomical barriers, as ventricular spaces, cranial bones, and the midline excluding for the region of the corpus callosum. An isotropic margin of 5mm was added around to obtain the Planning Target Volume (PTV-1). Radiotherapy treatment (RT) was delivered with a Linear Accelerator 6–10 MeV beam and 3D-Conformal or Intensity Modulated techniques up to a planned total dose of 60 Gy and with a standard fractionation (2Gy/day for 5 days per week). All patients received also temozolomide (TMZ), concurrently administered *per os* during RT, according to Stupp’s protocol (daily TMZ 75mg/m2 during the RT course, for 6 weeks followed by the sequential TMZ schedule (150–200mg/m2 for 5 days every 28 days) until disease progression (PD) or complete response (CR) after 12 cycles. After the completion of RT and concurrent TMZ administration, patients entered a scheduled follow-up program. Brain MRI scans were repeated at 4 weeks, 12–16 weeks, and then every 6 months or in any case showing clinical signs suggesting progressive disease (PD). Taking into account the fact that no patient of this series received antiangiogenic treatment, PD after RT-TMZ treatment was assessed using the RANO Criteria [19]. A diagnosis of pseudoprogression was made in cases showing an increase in tumor size and/or T1-contrast enhancement within 3– 6 months after the end of concomitant RT-TMZ, without worsening of neurological status and with stabilization or resolution in subsequent further MRIs studies. Imaging findings suggestive of radionecrosis were recorded. All the MRI examinations were revised for the compilation of this paper by a neuroradiologist (LEA). General and neurological examinations and blood counts and chemistry were obtained every three months.

### Statistical Analysis

Statistical analysis was performed using Statistical Package for Social Sciences (SPSS Inc, Chicago IL, USA). Kaplan-Meier survival curves were done and differences in PFS and OS were tested for statistical significance using the log-rank test. Significance level was set at p < 0.05. Cutoff value for *MGMT* methylation status was obtained by plotting receiver operating characteristic (ROC) curve by plotting true positive (sensitivity) versus false positive (100-specificity) for investigation of diagnostic efficacy of MGMT by considering GBM versus NND. The area under the ROC curve (AUC) assessed the accuracy, hence: if equals 1 means accurate test; <1-0.8 a good test; <0.8 – 0.7 a fair test, <0.7 – 0.6 a poor test, while <0.5 as worthless test [20].

## Results

The current study was carried out on FFPE samples from 20 NND and 58 GBM cases, their full clinical data were summarized in Table (1). No significant level was reached when considering gender between the two groups (NND *versus* GBM), while significant level was reached when age of the two groups were considered, for both groups *IDH1* mutation and *MGMT* methylation were as represented in Table (2). By plotting the ROC curve to report the methylation status between investigated groups the best cutoff point (methylation percentage) that discriminates between them was 66%, hence those <66 were represented as low - moderate methylation while those ≥ 66 were highly methylated (Figure 3). Accordingly, all NND patients were highly methylated (100%) while 30 out of 58 (51.7%) GBM cases reported high *MGMT* methylation and the remaining were low-moderate methylation at significant level *P*<0.0001. For *IDH1* mutation, it was detected in 15 GBM cases (25.9%) while the remaining (43, 74.1%) reported *IDH* wild type, and all NND patients reported *IDH* wild type at significant level *P*=0.011, as shown in Table (2).

**Table (1):** Clinical and demographic data for studied cases.

| **Factors** | **Non-Neurooncological patients**<br>**(n=20)** | **GBM**<br>**(n=58)** |
| --- | --- | --- |
| **Age (Mean)** |  |  |
| < 60 years | 20 (100%) | 42 (72.4%) |
| ≥ 60 years | 0 (0%) | 16 (27.6%) |
| | $X^2 = 6.9, P = 0.008$ | |
| **Gender** |  |  |
| Male | 10 (50%) | 35 (60.3%) |
| Female | 10 (50%) | 22 (39.7%) |
| **Pathology** |  | GBM (Grade IV) |
| **ECGO** |  |  |
| < 2 |  | 19 (32.8%) |
| = 2 |  | 39 (67.2%) |
| **Tumor site** |  |  |
| Lt |  | 29 (50%) |
| Right |  | 23 (39.7%) |
| Multiple |  | 6 (10.3%) |
| **Tumor size** |  |  |
| < 5cm |  | 22 (37.9%) |
| ≥ 5cm |  | 36 (62.1%) |
| **Surgical intervention** |  |  |
| Biopsy |  | 36 (62.1%) |
| Resection |  |  |
| - Total |  | 19 (32.8%) |
| - Sub-total |  | 3 (5.2%) |

**Table (2):** Investigated *MGMT* methylation and *IDH1* mutation among studied groups.

| Investigated items | Non-Neurooncological patients<br>(n=20) | GBM<br>(n=58) |
| --- | --- | --- |
| *MGMT* methylation |  |  |
| < 66% (low methylation) | 0 (0%) | 28 (48.3%) |
| ≥ 66% (highly methylated) | 20 (100%) | 30 (51.7%) |
| | $X^2 = 15, P < 0.0001$ | |
| *IDH1* mutation |  |  |
| Wild type | 20 (100%) | 43 (74.1%) |
| Mutant type | 0 (0%) | 15 (25.9%) |
| | $X^2 = 6.4, P = 0.011$ | |

**Figure 3:**
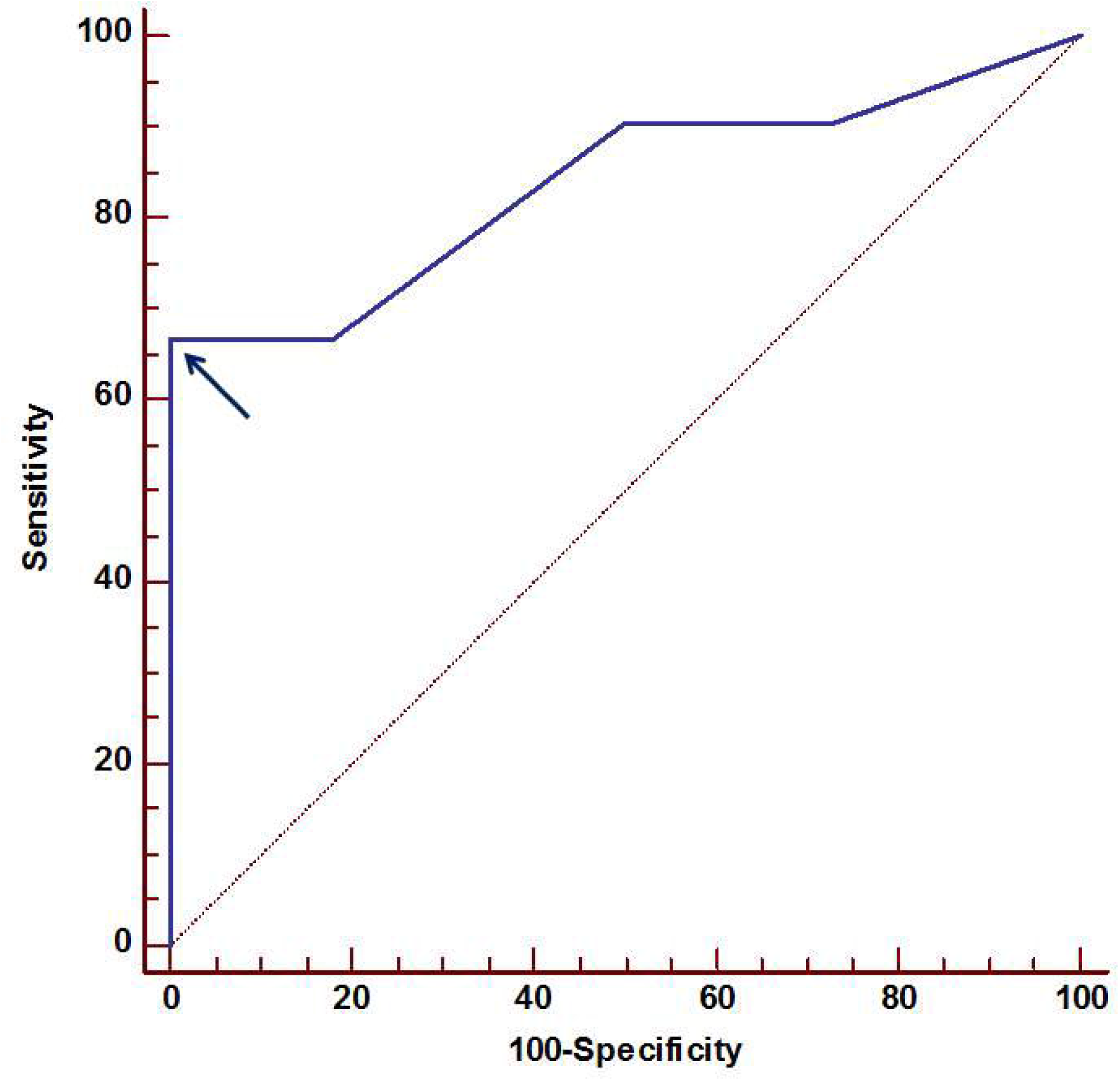
Receiver operating characteristic (ROC) curve for MGMT methylation among investigated groups. Arrow contributes to the best cutoff point that discriminates between high methylation (≥ 66%) versus low-moderate methylation at area under the curve (AUC)= 0.837, 95%CI= 0.723-0.917, at *P* = 0.0001.

Distributions of IDH1 mutation and *MGMT* methylation among GBM cases were presented in Table (3). For *IDH1* mutations significant levels were reported between *IDH1* mutation with both age, ECGO and surgical intervention, while for *MGMT* methylation was revealed significant with other factors apart from age and gender. Patients were treated with standard of care treatment protocol and patients were categorized according to their response to treatment as follows; complete response (CR), partial response (PR), stable disease (SD) and progressed disease (PD). Both *IDH1* mutation and *MGMT* methylation reported higher frequency among those patients with CR as reported in Table (4). When response of GBM patients were divided into either responders (CR, PR, SD) (n=30) *versus* non-responders (PD) (n=28) and GBM patients with both *IDH1* mutations and *MGMT* methylation were combined in one group (n=15) *versus* those GBM patients with either mutated, methylated or non in another group (n= 43), significant level was reached as all of GBM patients (15/15, 100%) with both *IDH1* mutated with *MGMT* methylated showed response to treatment as reported in Table (5).

**Table (3):** Distribution of *IDH1* mutation and *MGMT* methylation status among GBM cases.

| Factors | IDH1 Mutant | MGMT methylated ($\geq 66\%$) |
| --- | --- | --- |
| **Age (Mean)** |  |  |
| < 60 years | 0 (0%) | 24 (8%) |
| $\geq 60$ years | 15 (100%) | 6 (20%) |
| | $X^2 = 7.7, P = 0.005$ | |
| **Gender** |  |  |
| Male | 8 (53.3%) | 16 (53.3%) |
| Female | 7 (46.7) | 14 (46.7) |
| **ECGO** |  |  |
| < 2 | 11 (73.3%) | 19 (63.3%) |
| = 2 | 4 (26.7%) | 11 (36.7%) |
| | $X^2 = 15, P < 0.0001$ | $X^2 = 26, P < 0.0001$ |
| **Tumor site** |  |  |
| Lt | 8 (53.3%) | 17 (56.7%) |
| Right | 7 (46.7) | 13 (43.3%) |
| Multiple | 0 (0%) | 0 (0%) |
| | | $X^2 = 7.2, P = 0.027$ |
| **Tumor size** |  |  |
| < 5cm | 7 (46.7) | 17 (56.7%) |
| $\geq 5$cm | 8 (53.3%) | 13 (43.3%) |
| | | $X^2 = 9.3, P = 0.002$ |
| **Surgical intervention** |  |  |
| Biopsy | 8 (53.3%) | 13 (43.3%) |
| Resection |  |  |
| - Total | 4 (26.7%) | 14 (46.7%) |
| - Sub-total | 3 (20%) | 3 (10%) |
| | $X^2 = 9, P = 0.011$ | $X^2 = 9.98, P = 0.007$ |

**Table (4):**
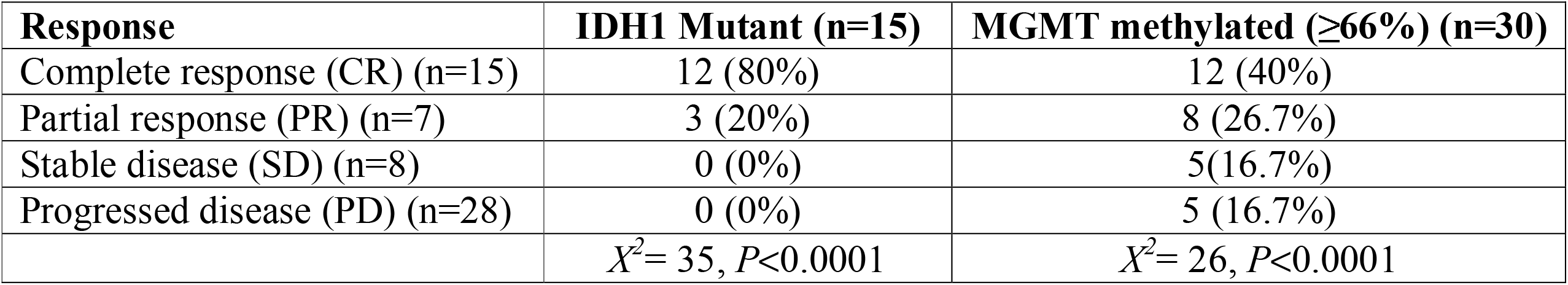
relation between response to treatment and investigated markers.

| **Response** | **IDH1 Mutant (n=15)** | **MGMT methylated ($\geq 66\%$) (n=30)** |
| --- | --- | --- |
| Complete response (CR) (n=15) | 12 (80%) | 12 (40%) |
| Partial response (PR) (n=7) | 3 (20%) | 8 (26.7%) |
| Stable disease (SD) (n=8) | 0 (0%) | 5 (16.7%) |
| Progressed disease (PD) (n=28) | 0 (0%) | 5 (16.7%) |
| | $X^2 = 35, P < 0.0001$ | $X^2 = 26, P < 0.0001$ |

**Table (5):** Distribution of the response of GBM patients when IDH1 mutation and MGMT methylation were combined.

| Response | Either *IDH1* mutated or MGMT methylated or both are not detected<br>(n=43) | *IDH1* mutation with *MGMT* methylation<br>(n=15) |
| --- | --- | --- |
| Responders<br>(n=30) | 15 (34.9%) | 15 (100%) |
| Non-responders<br>(n=28) | 28 (65.1%) | 0 (0%) |
| Statistics | $\chi^2 = 18.89, P < 0.0001$ | |

GBM patients were followed up for a median of 10 months and the estimated progression free survival (PFS) was 13 months while median overall survival (OS) was 16 months. Relation between survival pattern and estimated markers reported significant difference for *IDH1* mutation with PFS (log rank *X2* =9.2, *P*=0.002) and OS (log rank *X2* =8.99, *P*=0.003), as GBM patients reported to have IDH1 mutations revealed better PS and OS, similarly, *MGMT* methylation reported significant with PFS (log rank *X2* =17, *P*=0.0001) and OS (log rank *X2* =27, *P*=0.0001) as GBM patients with methylated MGMT showed better PFS and OS as reported in Figure (4 A-D). Moreover, survival pattern for patients with *IDH1* mutation with *MGMT* methylation was better (mean PFS =20 months, mean OS 26 months) than patients with either *IDH1* mutation or *MGMT* methylation alone (mean PFS =10 months, mean OS=15 months) as plotted in Figure (5A-B).

**Figure 4:**
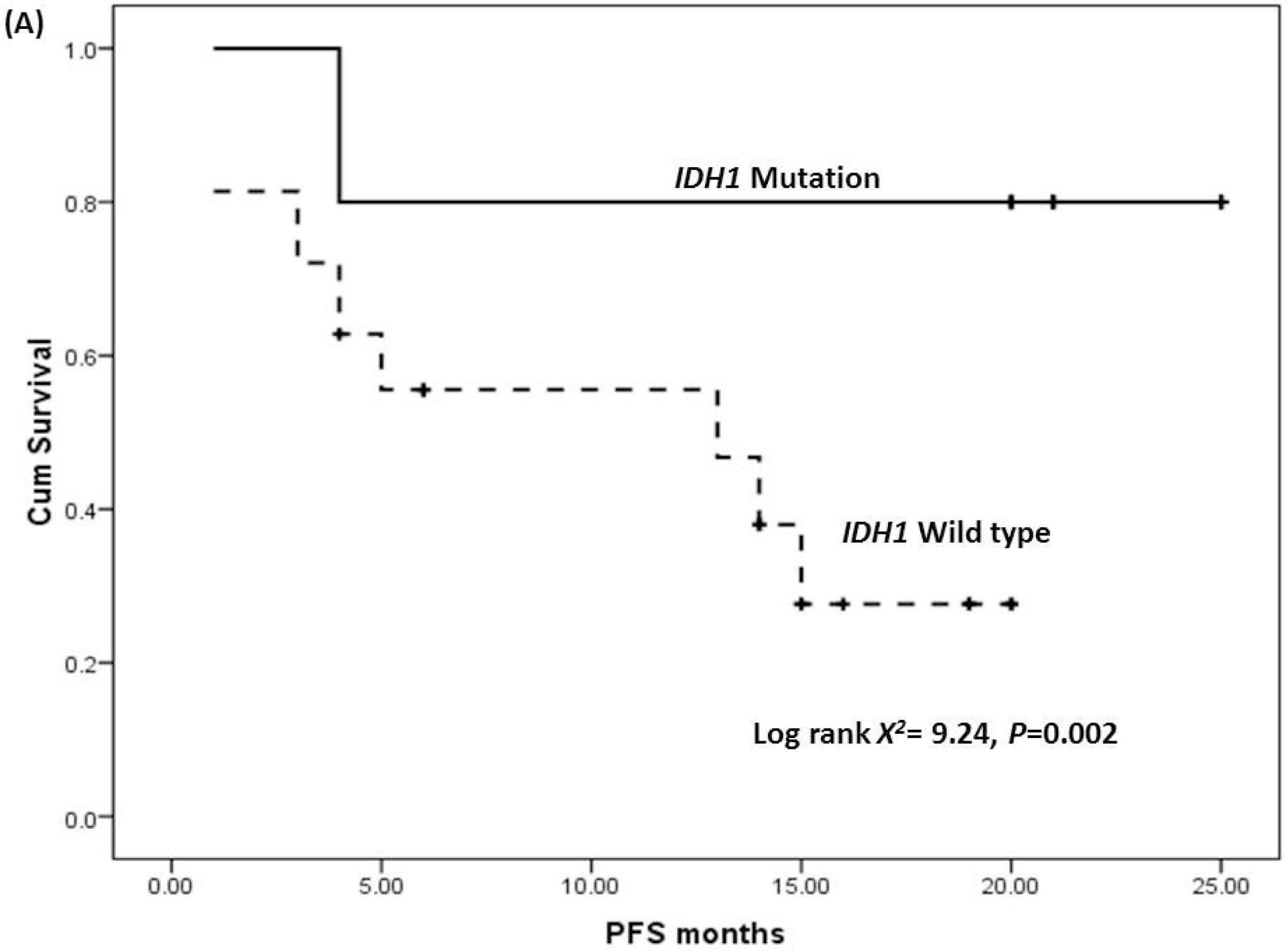
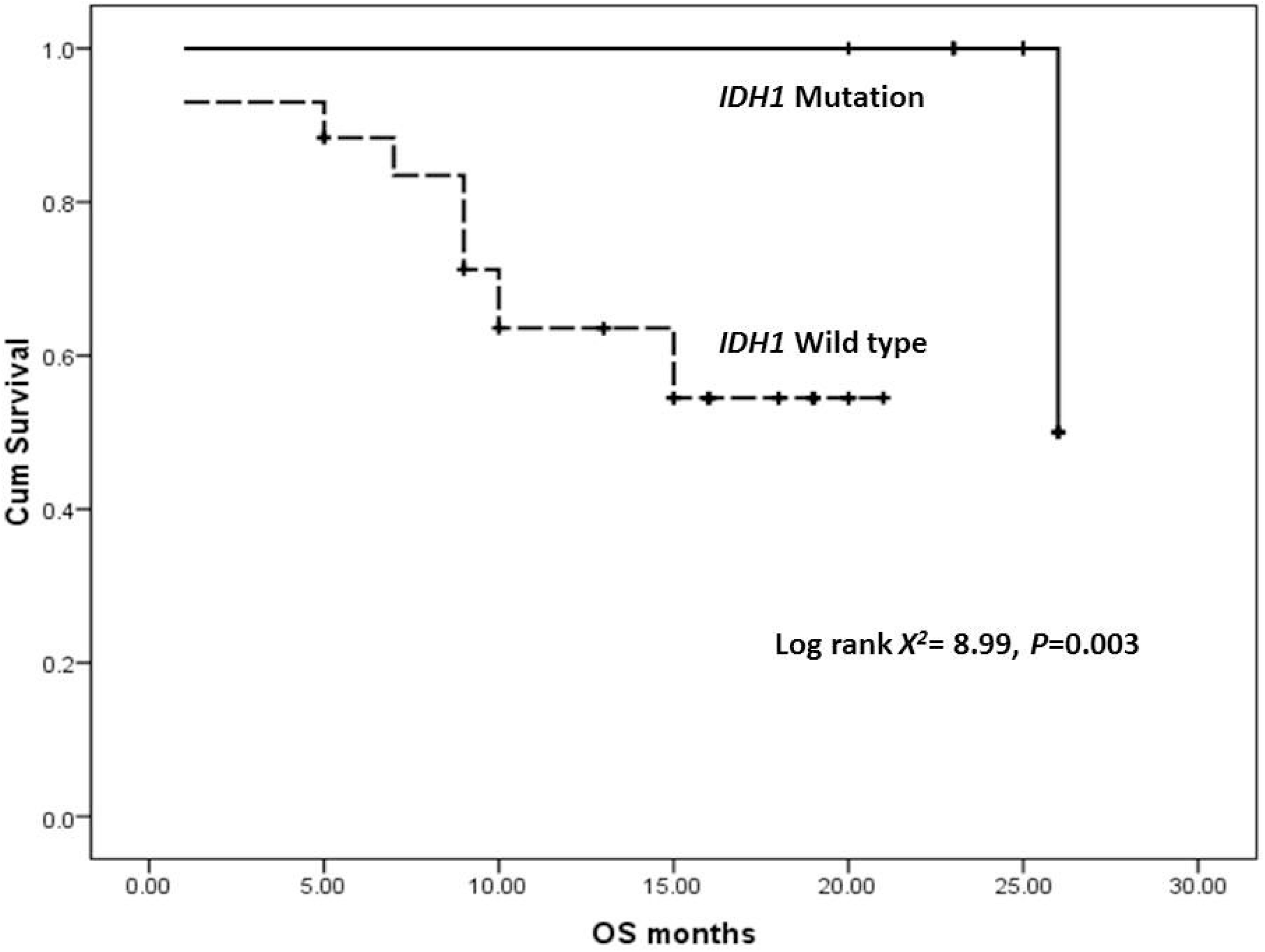
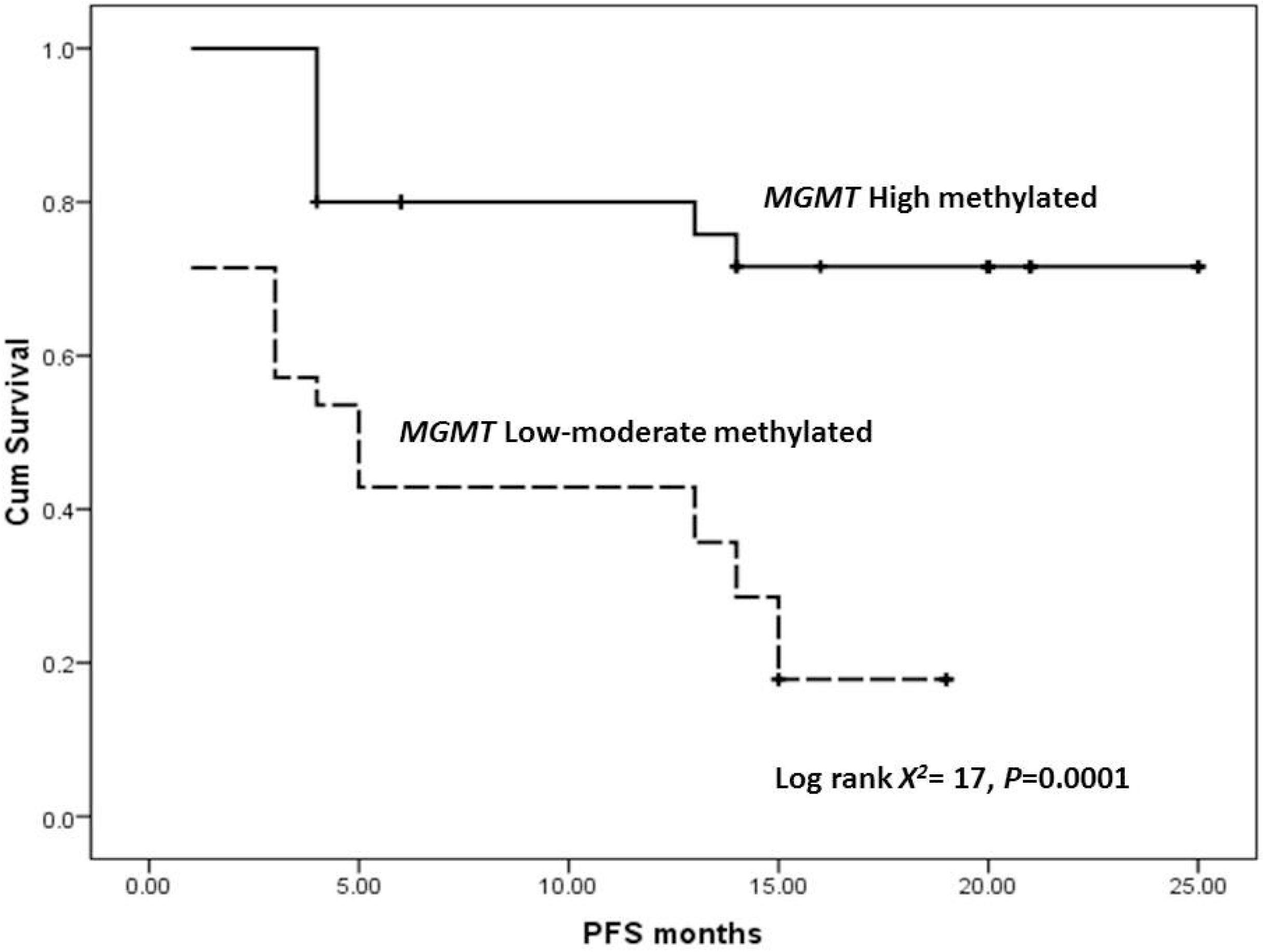
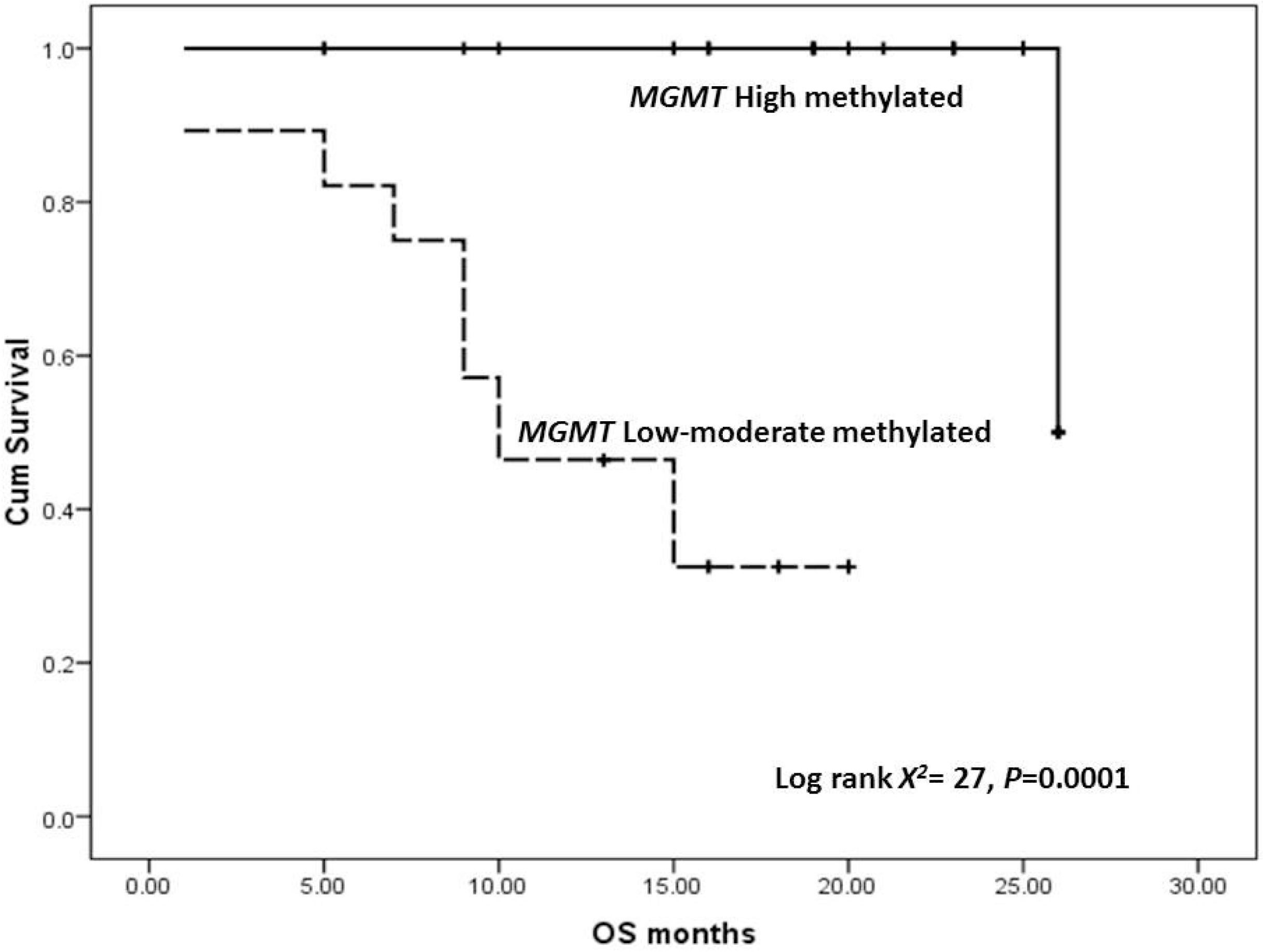
A) PFS for IDH1 mutation, B) OS for IDH1 mutation, C) PFS for MGMT methylation, D) OS for MGMT methylation.

**Figure 5:**
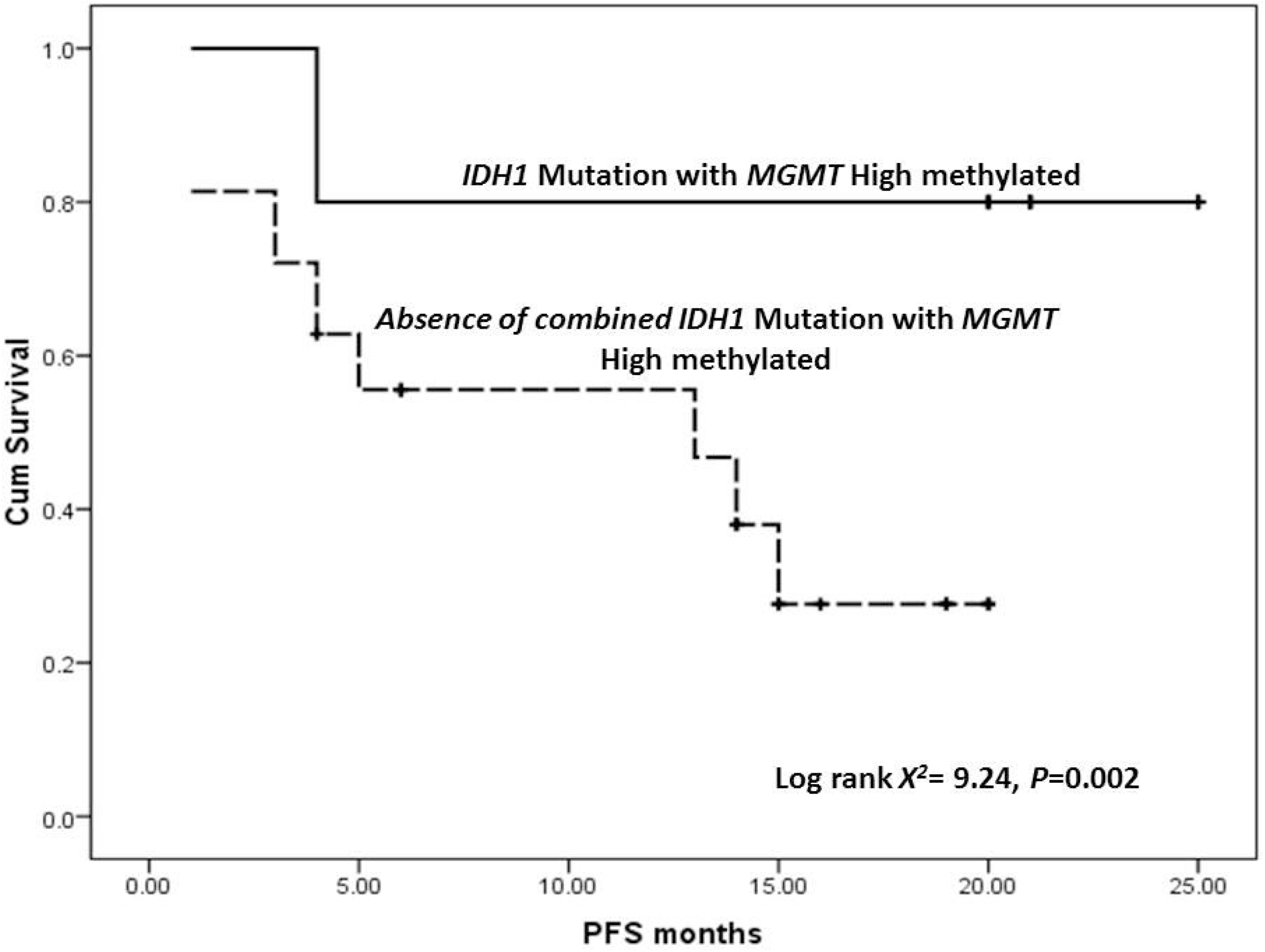
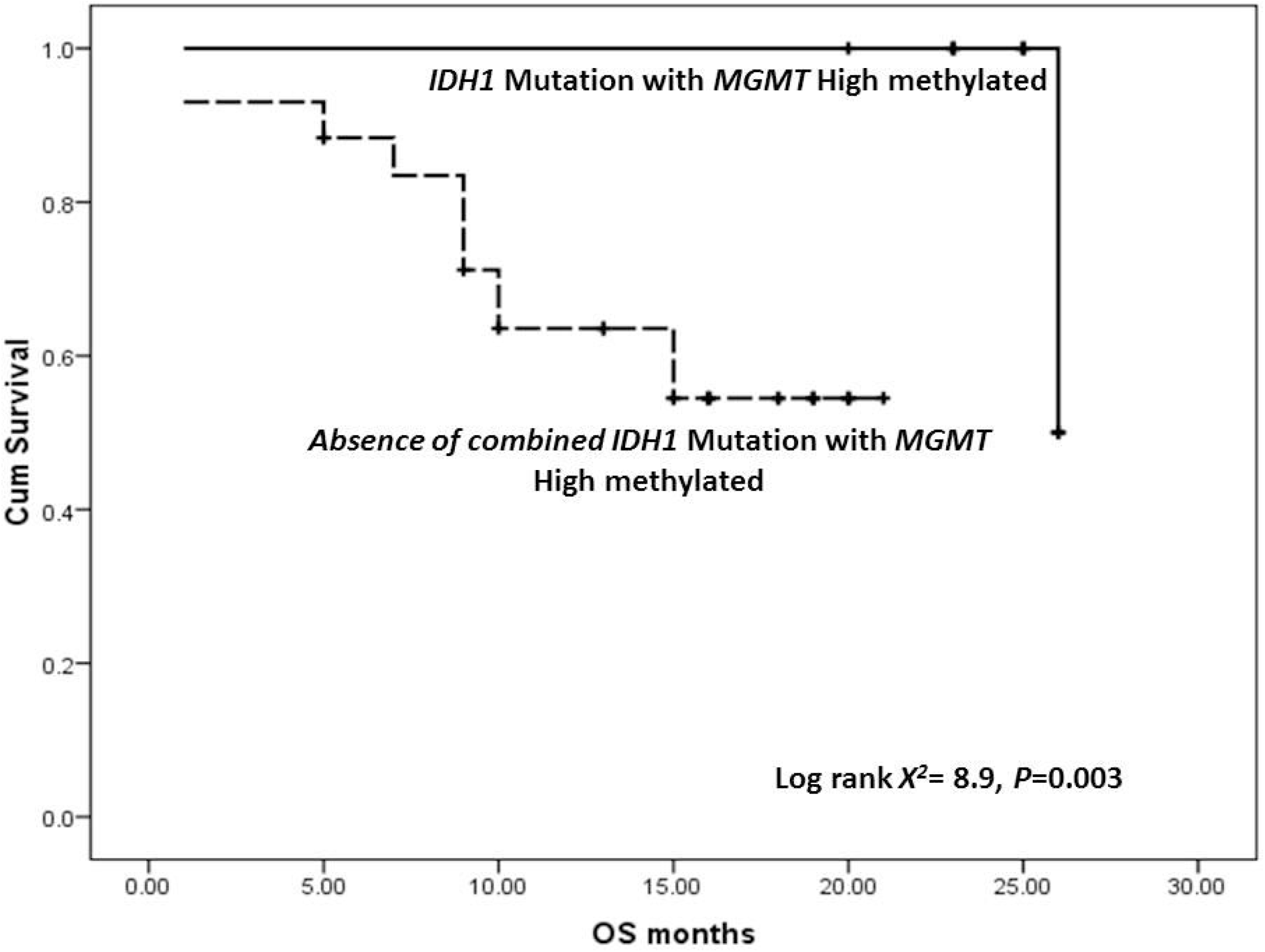
A) PFS or IDH1 mutation with MGMT methylation, B) OS for IDH1 mutation with MGMT methylation.

## Discussion

Alteration of many genes have been found to be implicated in pathogenesis of GBM, hence they may play an important role in predicting prognosis and response to treatment strategies [21]. In the current study, the impact of *IDH1* gene mutation and methylation of *MGMT promoter* were investigated among Egyptian GBM patients as compared to a group of NND. Among the investigated groups no significant difference was reported between their genders however significant difference was reported among their ages as all NND were below 60 years. This result emphasizes the relation between the increase of GBM among elderly which agree with previous reported studies [22,23] which may be attributed to the fact that aging may gradually suppresses immunosurveillance hence participates in GBM cell initiation and/or development [22].

Sanger sequencing is considered the “gold standard” for detection of *IDH1* mutations because of its high specificity and low false positive results but with some drawbacks as low sensitivity, consumes time and high-quality tissue samples to perform the reaction in addition needs manual interpretation [24]. Due to its significant to detect the occurrence of *IDH1* mutations in a fast method thus patients can gain the advantage from targeted therapies, so authors detected *IDH1* mutation using TaqMan™ competitive allele-specific probes (castPCR™) which has high sensitivity over Sanger sequencing (0.1% *versus* 10-25%, respectively) [25] and high specificity as minimal amounts of mutated DNA in the sample that have large quantities of normal wild-type DNA [17] since this technique uses oligonucleotides for the mutated allele so as to repress the normal allele [26]. Accordingly, in the current study*IDH1* mutation was not detect among patients with NND 0 out of 20 individuals (0%), the results of this study were agreed with earlier reported data [27] who reported that detection of *IDH1* mutation points to the existence of glioma and it cannot be ascribed to non-neoplastic diseases. For GBM cases *IDH1* mutation was detected in 15 out of 58 (25.9%). These results are in concordance with Kalkan and his colleagues [28] who reported the presence of *IDH1* mutations in 12.5% primary GBM cases which reveal that it is an early consequence in tumor genesis and this due to the fact that *IDH1* mutation reduce the action of NADPH which is important for cellular protection against oxidative stress giving rise to tumor genesis due to oxidative DNA damage [29].

Methylation status of *MGMT* is among the most studied molecular biomarkers in neuro-oncology because of its influence in therapeutic management of glioblastoma, thus its detection has been reported using different techniques [30]. However, debate remains about the most appropriate technique to be used, in the current study authors assessed methylation status using restriction enzyme that cut the unmethylated regions and hence the detected will be the methylated (REF). Although it was previously reported in several neuro-oncological centers as 10% as the biological cutoff [31], others reported that precise cutoff value might reflect their response to treatment [32]. In the current study as for the first time NND were included, the ROC was plotted between both groups as considering NND as reference (control) group hence the best cutoff point was 66% methylation (<66% as low-moderate methylation, ≥ 66% as highly methylated). By using this methylation cutoff, currently studied groups reported all NND patients with high *MGMT* methylation as compared to GBM cases as 51.9% were high *MGMT* methylation. Methylation of NND patients could be attributed to the previously reported findings of Teuber-Hanselmann and his colleagues that hypermethylation of *MGMT* arises in chronic neurological diseases that are not strictly associated to distinctive oncogenic viruses, pathogens, or neoplasms but that result in destruction of the myelin sheath by several ways [33].

Among the GBM cases; those reported *IDH1* mutation were of younger age (less than 60 years) than those with older ages; these results agreed with previously reported study by Kalkan and his colleagues [28], for *MGMT* methylation; significant levels were reached with factors like tumor size and tumor location which agreed with previous reports [34,35] as GBM patients with tumor size less than 5cm reported high methylation than others with mass more than 5cm, moreover it is generally recognized the site of tumor, as significant image characteristic related to genetic criteria, is associated with prognosis of patients [35]. Also, both *IDH1* mutation and *MGMT* methylation were reported at significant levels in GBM patients with ECGO < 2 which may indicate their usefulness as prognostication markers among GBM patients.

After patients were treated with standard of care treatment strategy, they were followed-up for median 10 months, GBM patients with *IDH1* mutations reported better PFS and OS than those with *IDH1* wild type. A finding that agreed with previously reported study [28] that *IDH1* mutations can be used as a prognostic marker for primary GBM patients since it is primary event in tumorigenesis. Regarding GBM patients with MGMT high methylation reported better PFD and OS as compared to those with low-moderate methylation, these results in concordance with [32].When GBM patients with both *IDH1* mutations and *MGMT* high methylation were considered, our results emphasized best PFS (20 months) and OS (25 months) indicating that detection of *IDH1* mutation combined with *MGMT* methylation is a better prognostic marker and estimates response of GBM patients to treatment than any of them alone this was agreed with previously reported finding [36] thus using both combined markers for predicting response to treatment and predicting survival pattern is obviously advised than using any of them alone.

## Conclusion

Current study reported the superiority of combined detection of *MGMT* methylation and IDH1 mutation among GBM as predictive and prognostic markers than using either of them alone. In addition, the method used for *MGMT* methylation and *IDH1* mutation detection reported to be highly sensitive than previously reported techniques.

## Data Availability

Availability of data and material: no data available to be shared.

## Abbreviation list

ARMS: Amplification Refractory Mutation System
*BRCA*: Breast cancer gene
CTV: Clinical Target Volume
CR: complete response
CT: Computerized tomography
DNA: deoxyribonucleic acid
ECOG: Ester Clinical Oncology Group
EGFR: epidermal growth factor receptor
GBM: glioblastoma multiforme
HE: Hematoxylin-eosin
*IDH*: isocitrate dehydrogenase
LOD: limit of detection
MRI: Magnetic resonance imaging
NND: non-neurooncological diseases
*MGMT*: O6-methylguanine-DNAmethyltransferase
PTV-1: planning Target Volume

## Declarations

### Ethics approval and consent to participate

Ethical approval of the study was obtained from the Medical Ethical Committee of National Research Centre (ID#20110), Egypt, and all participants signed their informed consent.

### Consent for publication

All authors agreed for sending the paper for publication.

### Availability of data and material

no data available to be shared.

### Competing interests

Authors declare no competing of interest.

### Funding

This paper is based upon work supported by Science, Technology & Innovation Funding Authority (STDF) Basic and Applied Research Support Grant Project (BARG) [No.25562]. The instruments listed in the current study were purchased through a grant from Science Technology & Innovation Funding Authority (STDF) through Capacity Building Grant Fund (CBG) [No. 4940], Egypt.

### Authors’ contributions

Study conception and design: MSM and MS. Provision of samples and clinical follow-up: LEA and MM. Acquisition of data: AR, NB, MH and AMN. Analysis and interpretation of data: MSM, MS; LEA, MKK, MM and MS. Drafting of manuscript: MS; LEA; MH and AR. Critical revision: all authors.

## Notes

### Competing Interest Statement

The authors have declared no competing interest.

### Author Declarations

After getting ethical approval from Medical Ethical Committee of National Research Centre (ID#20110)

## References

1 Pandith AA, Qasim I, Baba SM, Koul A, Zahoor W, Afroze D, Lateef A, Manzoor U, Bhat IA, Sanadhya D, Bhat AR, Ramzan AU, Mohammad F, Anwar I.(2020) Favorable role of IDH1/2 mutations aided with MGMT promoter gene methylation in the outcome of patients with malignant glioma. Future Sci OA. 9;7(3):FSO663. https://doi.org/10.2144/fsoa-2020-0057.

2 Stupp R, Hegi ME, Mason WP, van den Bent MJ, Taphoorn MJ, Janzer RC, Ludwin SK, Allgeier A, Fisher B, Belanger K, Hau P, Brandes AA, Gijtenbeek J, Marosi C, Vecht CJ, Mokhtari K, Wesseling P, Villa S, Eisenhauer E, Gorlia T, Weller M, Lacombe D, Cairncross JG, Mirimanoff RO; European Organisation for Research and Treatment of Cancer Brain Tumour and Radiation Oncology Groups; National Cancer Institute of Canada Clinical Trials Group.(2009) Effects of radiotherapy with concomitant and adjuvant temozolomide versus radiotherapy alone on survival in glioblastoma in a randomised phase III study: 5-year analysis of the EORTC-NCIC trial. Lancet Oncol. 10(5):459–66. https://doi.org/10.1016/S1470-2045(09)70025-7. Epub 2009 Mar 9.

3 Stupp R, Mason WP, van den Bent MJ, Weller M, Fisher B, Taphoorn MJ, Belanger K, Brandes AA, Marosi C, Bogdahn U, Curschmann J, Janzer RC, Ludwin SK, Gorlia T, Allgeier A, Lacombe D, Cairncross JG, Eisenhauer E, Mirimanoff RO; European Organisation for Research and Treatment of Cancer Brain Tumor and Radiotherapy Groups; National Cancer Institute of Canada Clinical Trials Group. (2005) Radiotherapy plus concomitant and adjuvant temozolomide for glioblastoma. N Engl J Med. 352(10):987–96. https://doi.org/10.1056/NEJMoa043330.

4 Pojo M, Costa BM, (2011) Molecular hallmarks of gliomas. In: Garami, M. (Ed.), Molecular Targets of CNS Tumors. In Tech, pp. 177–200.

5 Bleeker FE, Molenaar RJ, Leenstra S. (2012) Recent advances in the molecular understanding of glioblastoma. J Neurooncol. 108(1):11–27.

6 Zhang Y, Dube C, Gibert M Jr, Cruickshanks N, Wang B, Coughlan M, Yang Y, Setiady I, Deveau C, Saoud K, Grello C, Oxford M, Yuan F, Abounader R. (2018) The p53 Pathway in Glioblastoma. Cancers (Basel). 10(9):297. https://doi.org/10.3390/cancers10090297.

7 Fontanilles M, Marguet F, Ruminy P, Basset C, Noel A, Beaussire L, Viennot M, Viailly PJ, Cassinari K, Chambon P, Richard D, Alexandru C, Tennevet I, Langlois O, Di Fiore F, Laquerrière A, Clatot F, Sarafan-Vasseur N. (2020) Simultaneous detection of EGFR amplification and EGFRvIII variant using digital PCR-based method in glioblastoma. Acta Neuropathol Commun. 8(1):52. https://doi.org/10.1186/s40478-020-00917-6.

8 Yang JM, Schiapparelli P, Nguyen HN, Igarashi A, Zhang Q, Abbadi S, Amzel LM, Sesaki H, Quiñones-Hinojosa A, Iijima M.(2017) Characterization of PTEN mutations in brain cancer reveals that pten mono-ubiquitination promotes protein stability and nuclear localization. Oncogene. 36(26):3673–3685. https://doi.org/10.1038/onc.2016.493. Epub 2017 Mar 6.

9 Nageeb AM, Mohamed MM, Ezz El Arab LR, Khalifa MK, Swellam M. (2020) Next generation sequencing of BRCA genes in glioblastoma multiform Egyptian patients: a pilot study. Arch Physiol Biochem. 26:1–9. https://doi.org/10.1080/13813455.2020.1729814. Epub ahead of print.

10 Bleeker FE, Atai NA, Lamba S, Jonker A, Rijkeboer D, Bosch KS, Tigchelaar W, Troost D, Vandertop WP, Bardelli A, Van Noorden CJ. (2010)The prognostic IDH1(R132) mutation is associated with reduced NADP+-dependent IDH activity in glioblastoma. Acta Neuropathol. 119(4):487–94. https://doi.org/10.1007/s00401-010-0645-6. Epub 2010 Feb 4.

11 Romani M, Pistillo MP, Banelli B.(2018) Epigenetic Targeting of Glioblastoma. Front Oncol. 8: 448. https://doi.org/10.3389/fonc.2018.00448.

12 Swellam M, Ramadan A, Mahmoud MS, Hashim M, Emam M. (2017) Diagnostic role of aberrant DNA promoter methylation in ovarian cancer. Annual Research and Review in Biology 19(5):1–13.,ARRB.37658. DOI: 10.9734/ARRB/2017/37658

13 Ramadan A, Hashim M, Abouzid A, Swellam M. (2021) Clinical impact of PTEN methylation status as a prognostic marker for breast cancer. J Genet Eng Biotechnol. 19(1):66. https://doi.org/10.1186/s43141-021-00169-4.

14 Yu W, Zhang L, Wei Q, Shao A. (2020) O6-Methylguanine-DNA Methyltransferase (MGMT): Challenges and New Opportunities in Glioma Chemotherapy. Front Oncol. 9:1547. https://doi.org/10.3389/fonc.2019.01547.

15 Kleihues P. Cavenee W.K. eds. Pathology and Genetics of Tumours of the Central Nervous System (WHO Classification of Tumours), 2nd edition 1–314, IARC Lyon 02000.

16 Šestáková Š, Šálek C, Remešová H.(2019) DNA Methylation Validation Methods: a Coherent Review with Practical Comparison. Biol Proced Online. 21:19. https://doi.org/10.1186/s12575-019-0107-z.

17 Barbano R, Pasculli B, Coco M, Fontana A, Copetti M, Rendina M, Valori VM, Graziano P, Maiello E, Fazio VM, Parrella P. (2015) Competitive allele-specific TaqMan PCR (Cast-PCR) is a sensitive, specific and fast method for BRAF V600 mutation detection in Melanoma patients. Sci Rep. 5:18592. https://doi.org/10.1038/srep18592.

18 Olarte I, García A, Ramos C, Arratia B, Centeno F, Paredes J, Rozen E, Kassack J, Collazo J, Martínez A. (2019) Detection Of Mutations In The Isocitrate Dehydrogenase Genes (IDH1/IDH2) Using castPCRTM In Patients With AML And Their Clinical Impact In Mexico City. Onco Targets Ther. 12:8023-8031. 10.2147/OTT.S219703.

19 Uhm, J. (2010). “Updated response assessment criteria for high grade gliomas: response assessment in neuro-oncology working group,” Yearbook of Neurology and Neurosurgery, vol. 2010, pp. 118-119, 2010. group,” Yearbook of Neurology and Neurosurgery, vol. 2010, pp. 118–119,

20 Zweig MH, Campbell G, (1993) Receiver-operating characteristic (ROC) plots: a fundamental evaluation tool in clinical medicine. Clinical chemistry 39 (4): 561–577.

21 Zhang P, Xia Q, Liu L, Li S, Dong L. (2020) Current Opinion on Molecular Characterization for GBM Classification in Guiding Clinical Diagnosis, Prognosis, and Therapy. Front Mol Biosci. 7:562798. https://doi.org/10.3389/fmolb.2020.562798.

22 Ladomersky E, Scholtens DM, Kocherginsky M, Hibler EA, Bartom ET, Otto-Meyer S, Zhai L, Lauing KL, Choi J, Sosman JA, Wu JD, Zhang B, Lukas RV, Wainwright DA. (2019) The Coincidence Between Increasing Age, Immunosuppression, and the Incidence of Patients With Glioblastoma. Front Pharmacol. 10:200. https://doi.org/10.3389/fphar.2019.00200.

23 Minniti G, Lombardi G, Paolini S. (2019) Glioblastoma in Elderly Patients: Current Management and Future Perspectives. Cancers (Basel). 11(3):336. https://doi.org/10.3390/cancers11030336.

24 Gao J, Wu H, Wang L, Zhang H, Duan H, Lu J, Liang Z.(2016) Validation of targeted next-generation sequencing for RAS mutation detection in FFPE colorectal cancer tissues: comparison with Sanger sequencing and ARMS-Scorpion real-time PCR. BMJ Open. 6(1):e009532. https://doi.org/10.1136/bmjopen-2015-009532.

25 Huebner C, Weber R, Lloydd R. (2017) A HRM assay for identification of low level BRAF V600E and V600K mutations using the CADMA principle in FFPE specimens. Pathology. 49(7):776–783. https://doi.org/10.1016/j.pathol.2017.08.011.

26 Yang Y, Shen X, Li R, Shen J, Zhang H, Yu L, Liu B, Wang L. (2017) The detection and significance of EGFR and BRAF in cell-free DNA of peripheral blood in NSCLC. Oncotarget. 8(30):49773–49782. https://doi.org/10.18632/oncotarget.17937.

27 Horbinski C, Kofler J, Kelly LM, Murdoch GH, Nikiforova MN. (2009) Diagnostic use of IDH1/2 mutation analysis in routine clinical testing of formalin-fixed, paraffin-embedded glioma tissues. J Neuropathol Exp Neurol. 68(12):1319–25. https://doi.org/10.1097/NEN.0b013e3181c391be.

28 Kalkan R, Atli Eİ, Özdemir M, Çiftçi E, Aydin HE, Artan S, Arslantaş A. (2015) IDH1 mutations is prognostic marker for primary glioblastoma multiforme but MGMT hypermethylation is not prognostic for primary glioblastoma multiforme. Gene. 554(1):81–6. https://doi.org/10.1016/j.gene.2014.10.027. Epub 2014 Oct 14.

29 Lee SM, Koh HJ, Park DC, Song BJ, Huh TL, Park JW. (2002) Cytosolic NADP(+)-dependent isocitrate dehydrogenase status modulates oxidative damage to cells. Free Radic Biol Med. 32(11):1185–96. https://doi.org/10.1016/s0891-5849(02)00815-8.

30 Rosas-Alonso R, Colmenarejo-Fernandez J, Pernia O, Rodriguez-Antolín C, Esteban I, Ghanem I, Sanchez-Cabrero D, Losantos-Garcia I, Palacios-Zambrano S, Moreno-Bueno G, de Castro J, Martinez-Marin V, Ibanez-de-Caceres I. (2021) Clinical validation of a novel quantitative assay for the detection of MGMT methylation in glioblastoma patients. Clin Epigenetics. 13(1):52. https://doi.org/10.1186/s13148-021-01044-2.

31 Xie H, Tubbs R, Yang B. (2015) Detection of MGMT promoter methylation in glioblastoma using pyrosequencing. Int J Clin Exp Pathol. 8(2):1790-6. PMID: 25973069.

32 Radke J, Koch A, Pritsch F, Schumann E, Misch M, Hempt C, Lenz K, Löbel F, Paschereit F, Heppner FL, Vajkoczy P, Koll R, Onken J. (2019) Predictive MGMT status in a homogeneous cohort of IDH wildtype glioblastoma patients. Acta Neuropathol Commun. 7(1):89. https://doi.org/10.1186/s40478-019-0745-z. Erratum in: Acta Neuropathol Commun. 7(1):131.

33 Teuber-Hanselmann S, Worm K, Macha N, Junker A. (2021) MGMT-Methylation in Non-Neoplastic Diseases of the Central Nervous System. Int J Mol Sci. 22(8):3845. https://doi.org/10.3390/ijms22083845.

34 Iliadis G, Kotoula V, Chatzisotiriou A, Televantou D, Eleftheraki AG, Lambaki S, Misailidou D, Selviaridis P, Fountzilas G. (2012) Volumetric and MGMT parameters in glioblastoma patients: survival analysis. BMC Cancer. 12:3. https://doi.org/10.1186/1471-2407-12-3.

35 Han Y, Yan LF, Wang XB, Sun YZ, Zhang X, Liu ZC, Nan HY, Hu YC, Yang Y, Zhang J, Yu Y, Sun Q, Tian Q, Hu B, Xiao G, Wang W, Cui GB.(2018) Structural and advanced imaging in predicting MGMT promoter methylation of primary glioblastoma: a region of interest based analysis. BMC Cancer. 18(1):215. https://doi.org/10.1186/s12885-018-4114-2.

36 Molenaar RJ, Verbaan D, Lamba S, Zanon C, Jeuken JW, Boots-Sprenger SH, Wesseling P, Hulsebos TJ, Troost D, van Tilborg AA, Leenstra S, Vandertop WP, Bardelli A, van Noorden CJ, Bleeker FE. (2014) The combination of IDH1 mutations and MGMT methylation status predicts survival in glioblastoma better than either IDH1 or MGMT alone. Neuro Oncol. 16(9):1263–73. https://doi.org/10.1093/neuonc/nou005. Epub 2014 Feb 6.

